# Comparative assessment of multiple COVID-19 serological technologies supports continued evaluation of point-of-care lateral flow assays in hospital and community healthcare settings

**DOI:** 10.1101/2020.06.02.20120345

**Authors:** Suzanne Pickering, Gilberto Betancor, Rui Pedro Galão, Blair Merrick, Adrian W. Signell, Harry D. Wilson, Mark Tan Kia Ik, Jeffrey Seow, Carl Graham, Sam Acors, Neophytos Kouphou, Kathryn J.A. Steel, Oliver Hemmings, Amita Patel, Gaia Nebbia, Sam Douthwaite, Lorcan O’Connell, Jakub Luptak, Laura E. McCoy, Philip Brouwer, Marit J. van Gils, Rogier W. Sanders, Rocio Martinez Nunez, Karen Bisnauthsing, Geraldine O’Hara, Eithne MacMahon, Rahul Batra, Michael H. Malim, Stuart J.D. Neil, Katie J. Doores, Jonathan D. Edgeworth

## Abstract

There is a clear requirement for an accurate SARS-CoV-2 antibody test, both as a complement to existing diagnostic capabilities and for determining community seroprevalence. We therefore evaluated the performance of a variety of antibody testing technologies and their potential as diagnostic tools. A highly specific in-house ELISA was developed for the detection of anti-spike (S), -receptor binding domain (RBD) and -nucleocapsid (N) antibodies and used for the cross-comparison of ten commercial serological assays – a chemiluminescence-based platform, two ELISAs and seven colloidal gold lateral flow immunoassays (LFIAs) – on an identical panel of 110 SARS-CoV-2-positive samples and 50 pre-pandemic negatives. There was a wide variation in the performance of the different platforms, with specificity ranging from 82% to 100%, and overall sensitivity from 60.9% to 87.3%. However, the head-to-head comparison of multiple sero-diagnostic assays on identical sample sets revealed that performance is highly dependent on the time of sampling, with sensitivities of over 95% seen in several tests when assessing samples from more than 20 days post onset of symptoms. Furthermore, these analyses identified clear outlying samples that were negative in all tests, but were later shown to be from individuals with mildest disease presentation. Rigorous comparison of antibody testing platforms will inform the deployment of point-of-care technologies in healthcare settings and their use in the monitoring of SARS-CoV-2 infections.

## INTRODUCTION

As of the 1^st^ of June 2020, over 6 million cases of SARS-CoV-2 have been confirmed worldwide, accounting for more than 370,000 deaths (https://covid19.who.int/). Lack of treatments or vaccines have forced governments to adopt strict quarantine strategies in an attempt to control the spread of the virus, causing major economical disturbances as well as adversely affecting quality of life and healthcare provision.

Current guidelines by leading health bodies, including the Centers for Disease Control and Prevention (CDC) in the US and Public Health England (PHE) in the UK, recommend SARS-CoV-2 diagnosis from upper or lower respiratory specimens (including nasopharyngeal swabs and bronchoalveolar lavage) using real-time RT-PCR, typically targeting the nucleocapsid (N) or RNA-dependent RNA polymerase (RdRp) genes [1,2]. These tests are highly sensitive – capable of detecting vestigial viral RNA levels – and are optimal for the early detection of the virus. However, the performance of the test is dependent on the time the sample is collected, with viral load declining after the first week of symptoms [3,4].

There is, therefore, a clear requirement for accurate serology testing as a companion diagnostic to PCR-based testing. This is highlighted by the recent appearance of clusters of paediatric inflammatory multisystem syndrome (PIMS) and other hyperimmune reactions associated with SARS-CoV-2 infection [5,6]. Presentation is delayed relative to active viral infection, with the detection of antibody responses being key to clinical diagnosis. In addition, monitoring population seroprevalence will be central to future public health planning based on disease susceptibility and herd immunity [7]. For this to be meaningful, it is imperative that antibody detection methods are affordable, reliable, and readily accessible.

However, with no ‘gold standard’ for antibody testing and an incomplete knowledge of the immunology of COVID-19, evaluating tests with the assumption that antibodies ‘should’ be there, and comparatively to RT-PCR, is problematic. Head-to-head comparisons of multiple sero-diagnostic assays on identical samples therefore provides a robust assessment of individual assay performance. Accordingly, we developed a highly specific semi-quantitative ELISA for the detection of anti-spike (S), –S receptor binding domain (RBD) and –N antibodies, and used this to cross-evaluate ten commercial antibody tests (seven lateral flow immunoassays (LFIAs), one chemiluminescent assay and two ELISAs) on a collection of 110 serum samples from confirmed RNA positive patients, and 50 pre-pandemic samples from March 2019. Our results demonstrate a wide variation in the performance of the different platforms, ranging from 60.9% to 87.3% sensitivity and from 82% to 100% specificity. As expected, performance is highly dependent on the time the sample was taken post onset of symptoms (POS) and disease severity. Results obtained in this work have enabled the diagnostic-grade validation for one of the LFIAs evaluated for pilot clinical use for adult and paediatric patients with a range of clinically-suspected COVID-19 inflammatory syndromes at Guy’s and St Thomas’ Hospitals.

## RESULTS

### Study samples

110 serum samples collected from 87 individuals between the 4^th^ of March and 21^st^ of April 2020 at St Thomas’ Hospital were used to compare a panel of serological assays. At the time of study, UK government guidelines limited SARS-CoV-2 testing to individuals requiring hospitalisation, and all 87 individuals had RT-PCR-confirmed SARS-CoV-2 infection. Samples were representative of typical hospital admissions during the period, with a spectrum of clinical severities from mild (requiring no respiratory support) to critical (requiring extra-corporeal membrane oxygenation (ECMO)), and a range of time points after self-reported onset of symptoms (1 to 30 days) (**Table 1**).

**Table 1.** Baseline of patient cohort clinical characteristics.

| **Demographics** |  |
| --- | --- |
| Mean Age (years) | 58.2 +/- 16.6 |
| Female | 29 (33.3%) |
| **Level of Respiratory Support** |  |
| No support | 11 (12.6%) |
| Supplemental oxygen | 23 (26.4%) |
| Non- invasive ventilation | 1 (1.1%) |
| Mechanical ventilation | 46 (52.8%) |
| ECMO | 6 (6.9%) |
| **Disease Severity** |  |
| Level 0 | 11 (12.6%) |
| Level 1 | 15 (17.2%) |
| Level 2 | 4 (4.6%) |
| Level 3 | 3 (3.4%) |
| Level 4 | 48 (55.2%) |
| Level 5 | 6 (6.9%) |
| Died | 21 (24.1%) |
| **Comorbidities** |  |
| Hypertension | 40 (45.9%) |
| Obesity | 38 (43.7%) |
| T2DM | 26 (29.9%) |
| Hypercholesterolaemia | 10 (11.5%) |
| Chronic respiratory disease * | 12 (13.8%) |
| CKD | 5 (5.7%) |
| ESRF | 4 (4.6%) |
| Renal transplant | 3 (3.4%) |
| Hypothyroidism | 3 (3.4%) |
| T1DM | 2 (2.3%) |
| HIV | 1 (1.1%) |

\* COPD, interstitial lung disease, bronchiectasis, asthma

### In-house ELISA development

An in-house ELISA was developed to measure antibody responses against the full-length S, the RBD and N. Recombinant S and RBD were expressed in HEK 293F cells and purified by affinity and size exclusion chromatography. N was expressed in and purified from *E. coli*. Prepandemic serum samples from several cohorts were used to determine the lower limit of the assay, and samples from hospital patients with confirmed SARS-CoV-2 infection (from the described serum samples in **Table 1)** were used as positive controls. A total of 320 negative control samples included sera from individuals attending St Thomas’ Hospital in March 2019, sera from vaccination studies, healthy volunteers, and individuals with acute EBV infection.

All 20 serum samples from PCR positive ICU patients taken at least 10 days post symptoms showed strong IgG binding to S, RBD and N (**Figure 1A**). In contrast, although high IgM reactivity was also observed to S and RBD in some individuals (**Figure 1A**), only 5 of the 320 negative control samples showed IgG reactivity to S or RBD. High IgM and IgG reactivity was observed in the pre-pandemic samples against N (**Figure 1A**) suggestive of potential cross-reaction with seasonal coronaviruses. Of note, all individuals with acute EBV infection had high IgM reactivity to N and RBD. Importantly, none of the pre-COVID sera had detectable IgG binding to N and S or N and RBD (**Figure 1B**). Taking into account the reactivity of negative control samples in this ELISA, 100% specificity could be reached using a cut-off where IgG against N or S both have OD values at least 4-fold above the wells containing secondary antibody only (**Figure 1B**).

**Figure 1.**
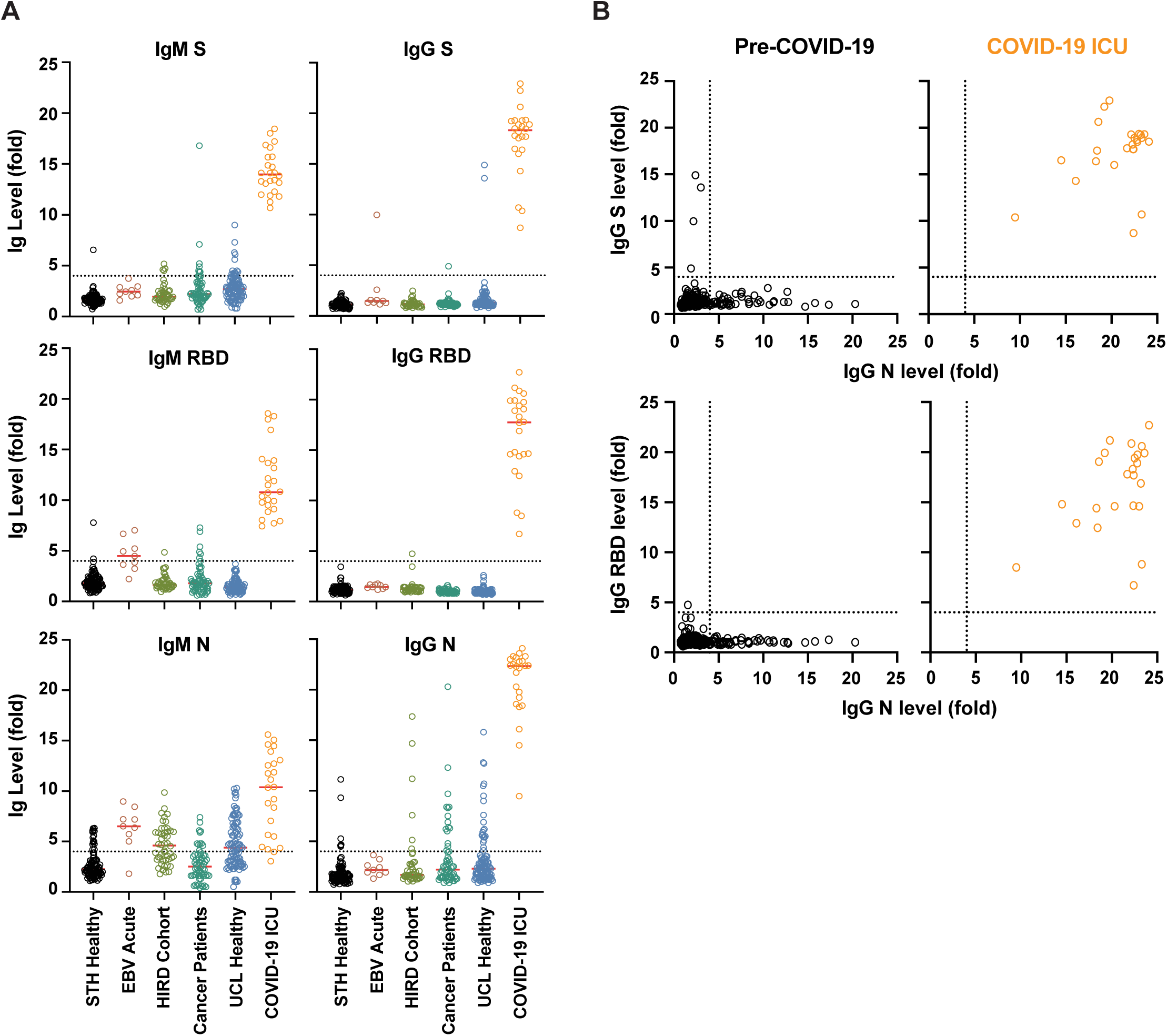
Validation of a SARS-CoV-2 ELISA for measuring IgG and IgM in Covid-19 patients. (**A**) > 300 pre-pandemic serum/plasma samples were assayed for IgM and IgG against SARSCoV-2 N, a stabilised and uncleaved S protein, and the RBD from S. Sera/plasma came from healthy volunteers (UCL healthy), vaccine trials (HIRD cohort [18]), cancer patients, and emergency admissions to St Thomas’ hospital in March 2019 (STH healthy). The IgM and IgG binding were compared to that of 24 SARS-CoV-2 PCR positive ICU patients collected in March and April 2020. Sera and plasma were diluted to 1:50 and 1:25 respectively. The values reported are the fold-change in OD above background. **(B)** Analysis of IgG and IgM binding of pre-Covid-19 human sera/plasma compared to sera from SARS-CoV-2 infected patients revealed that IgG against both N and S distinguished these two groups. The fold-change above background for IgG against N was plotted against the fold-change above background for S IgG binding and RBD IgG binding for each individual.

### Comparison of serological tests

All serological assays were evaluated with the same set of 110 serum samples from confirmed SARS-CoV-2-positive individuals. Each of the samples was tested: for anti-SARS-CoV-2 IgM by in-house ELISA and seven LFIAs; for IgA by commercial ELISA (**Figure 2**); and for IgG by inhouse ELISA, seven LFIAs, a commercial ELISA and for total antibody (IgG, IgM and IgA) using a chemiluminescent assay (**Figure 3**). The commercial ELISAs detect anti-S1 antibodies; for the LFIAs this is undisclosed proprietary information, but for some of the tests is known to be S. With no existing gold standard for the assessment of the serological response to SARS-CoV-2, we started by comparing commercial serological assays with the best performing configuration of the in-house ELISA (detection of anti-S IgM and IgG antibodies), which was also most likely to represent antibodies detected by the commercial tests. For the purpose of illustration, the intensity of bands shown in a positive LFIA test, or signal strength in commercial ELISA or chemiluminescent assay, is reproduced as a heatmap. However, the visible detection of a band or result above a given manufacturer’s threshold scored as a positive result regardless of classification. For samples giving a strong response by ELISA (> 9-fold), the majority of the commercial assays show a consensus positive result. Weaker antibody responses yielded mixed results from the LFIAs, with a clear pattern of increasing detection of antibodies seen across all tests with increasing time POS. Samples from days 1–9 gave an extremely mixed picture, indicative of an early evolving immune response. For the detection of IgM, 11 of the 72 samples from days 10 or more POS were negative by anti-S ELISA. 5 of these 11 samples were positive, in some cases only weakly, for at least two LFIAs and/or IgA. The remaining 6 were negative in all 7 LFIA tests and for IgA (indicated with a yellow circle in **Figure 2**). These same 6 samples were negative for IgG by in-house and commercial ELISA, in all 7 LFIAs and by chemiluminescent assay (indicated with a yellow circle in **Figure 3**). Importantly, all 6 samples came from individuals with a disease severity score of 0. Later time points were obtained for three of these individuals (both of the day 10 samples and the sixth day 14 sample) and the next available samples were found to be strongly positive for IgG and IgM by LFIA (days 47, 77 and 51 POS, respectively). Further investigation into the nature of the negative day 23 sample revealed that it was an error in self-reporting the time of symptom onset: this individual had COVID-19-compatible symptoms for 23 days prior to sampling, yet tested negative for RNA 10 days POS. 21 days POS the individual tested positive for RNA, therefore the more likely time of sampling was between 2 and 12 days POS. Samples were not re-classified, as they are representative of the real-time analyses being performed during the peak of a pandemic. However, the day 23 sample was omitted from sensitivity calculations.

**Figure 2.**
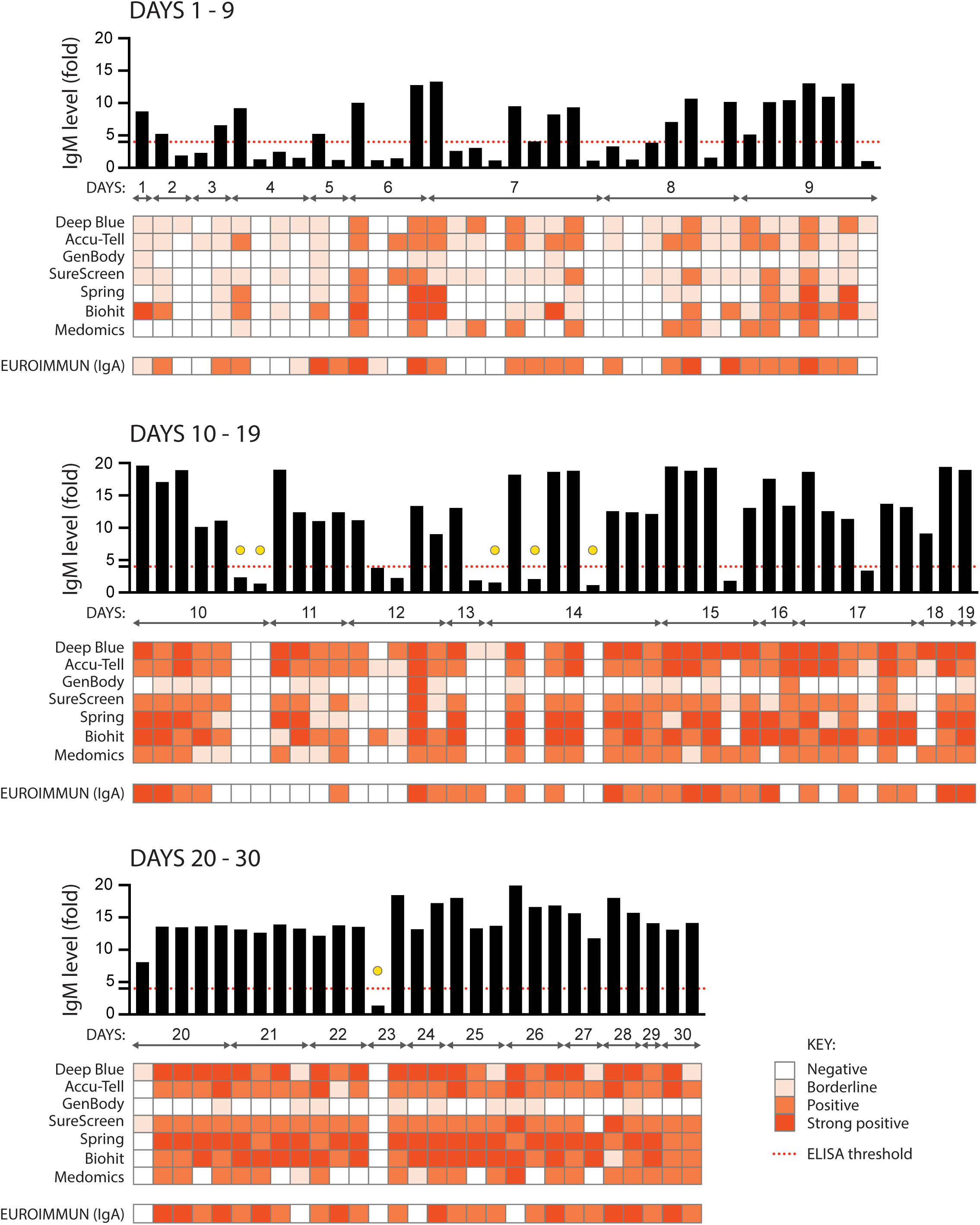
Comparison of nine serological assays for the detection of anti-SARS CoV-2 IgM and IgA. 110 serum samples from 87 individuals with confirmed SARS-CoV-2 infection (RNA+ by RTPCR) were assayed for anti-SARS CoV-2 IgM using an in-house anti-S ELISA (shown in the graph across the top of each panel, black bars), seven colloidal gold lateral flow tests (Deep Blue, Accu-Tell, GenBody, SureScreen, Spring, Biohit and Medomics), and for anti-S IgA using a commercial ELISA (EUROIMMUN). The threshold for a positive result in the in-house ELISA is set at 4-fold above background, as indicated by the red dashed line. Results for the other tests are represented as heatmaps, with colour intensity corresponding to strength of signal for each test. Samples are grouped according to days post onset of COVID-19 symptoms, and squares aligned in columns under each bar of the graph show results for a single serum sample. Yellow circles indicate samples from 10 days or more POS that were negative by ELISA and in at least 6 other tests, as detailed in the text.

**Figure 3.**
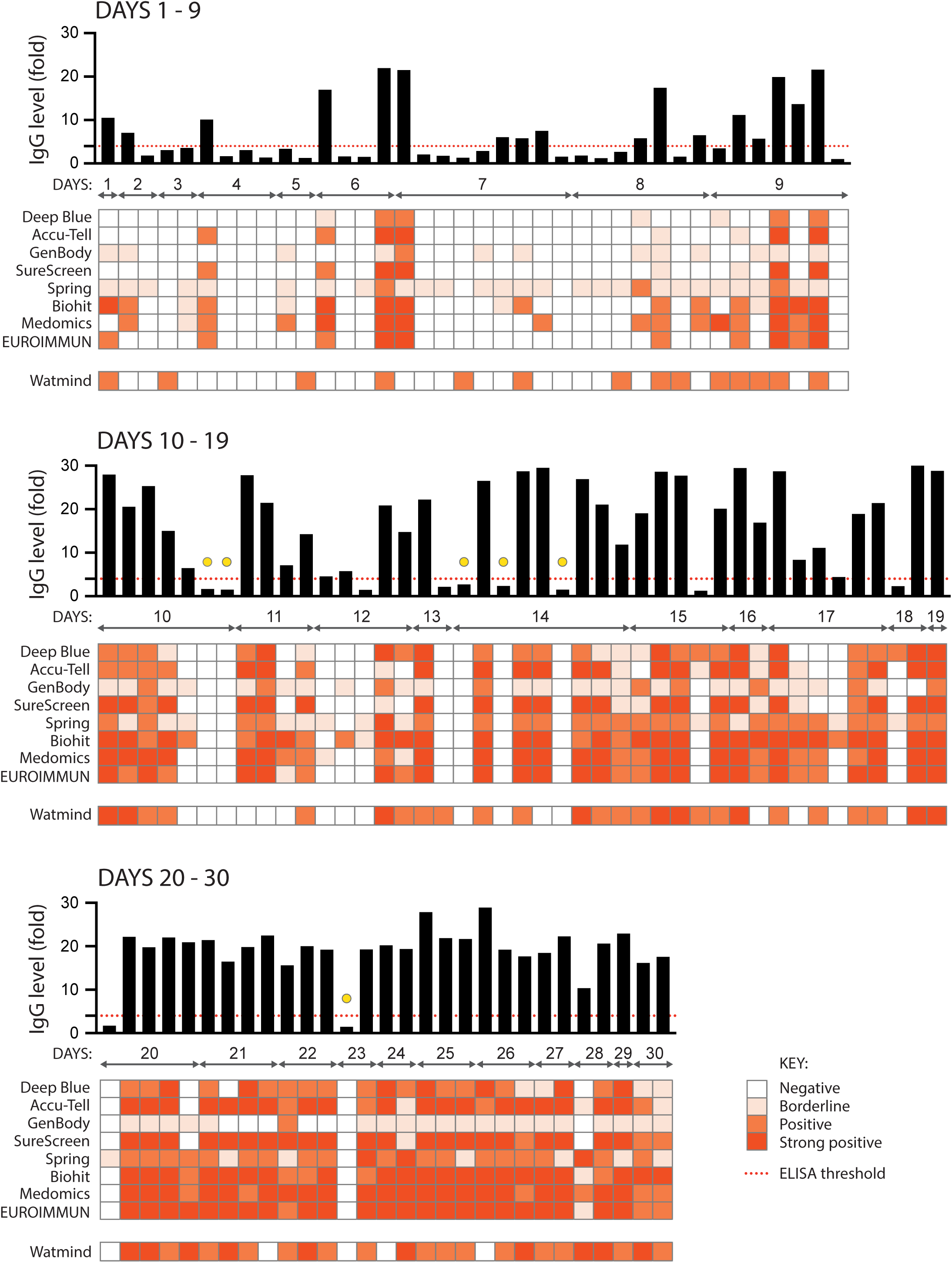
Comparison of ten serological assays for the detection of anti-SARS CoV-2 IgG. As for in **Figure 2**, the same 110 serum samples were assessed for the presence of anti-SARS CoV-2 IgG. Each sample was assayed using an in-house anti-S ELISA (shown in the graph across the top of each panel, black bars), seven colloidal gold lateral flow tests (Deep Blue, Accu-Tell, GenBody, SureScreen, Spring, Biohit and Medomics), and a commercial ELISA (EUROIMMUN). A chemiluminescent assay for total anti-SARS CoV-2 IgM, IgG and IgA (Watmind) was also included. The threshold for a positive result in the in-house ELISA is set at 4-fold above background, as indicated by the red dashed line.

Overall, with the exception of GenBody and Watmind, all tests gave cross-assay agreements between 81.8 and 95.4% (**Figure 4 and Supplementary Figure A**). The highest level of agreement was seen between the in-house ELISA, SureScreen, Accu-Tell, Spring and EUROIMMUN tests. Interestingly, the in-house ELISA IgM and EUROIMMUN IgA results showed particularly good agreement (94.5%), although the EUROIMMUN detected IgA more frequently in early samples compared with the IgM detected by in-house ELISA (**Figure 2**).

**Figure 4.**
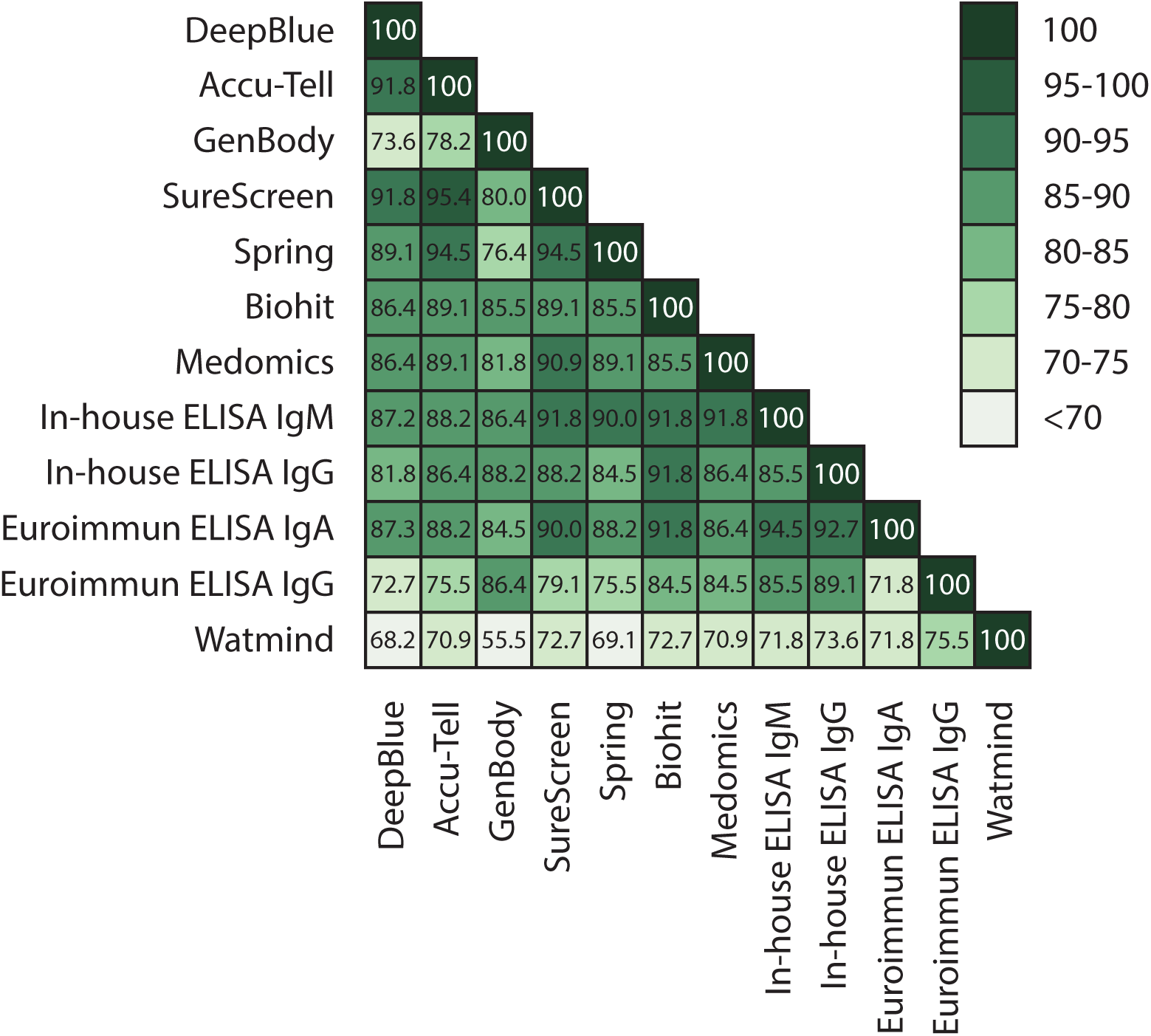
Comparative tables. Serological assays were compared and the percentage agreement between each of the samples in the assays is represented within each box.

### Assay specificity and sensitivity

Results from the 110 SARS-CoV-2-positive samples and a further 50 pre-pandemic negative samples were used to evaluate assay sensitivities and specificities (**Figure 5A, Supplementary Figure B and Supplementary Tables 2–4**). Cross comparison of overall specificities and sensitivities led to the shortlisting of six tests with the highest specificity and sensitivity (ELISA IgM and IgG, SureScreen, Accu-Tell, Spring and EUROIMMUN IgA). These were the same tests that gave the best agreement in the cross-assay comparisons. Notably, sensitivity increased for all tests with increasing days POS, with antibodies being variably detected at early times (**Figure 5B**). Deep Blue, Accu-Tell, SureScreen and Spring also displayed the highest levels of sensitivity at less than 10 days. In sum, Deep Blue, Accu-Tell, SureScreen, Spring, Biohit, Medomics, EUROIMMUN (IgA and IgG) and in-house ELISAs (IgM and IgG) all had sensitivities above 95% for samples taken ³20 days POS.

**Figure 5.**
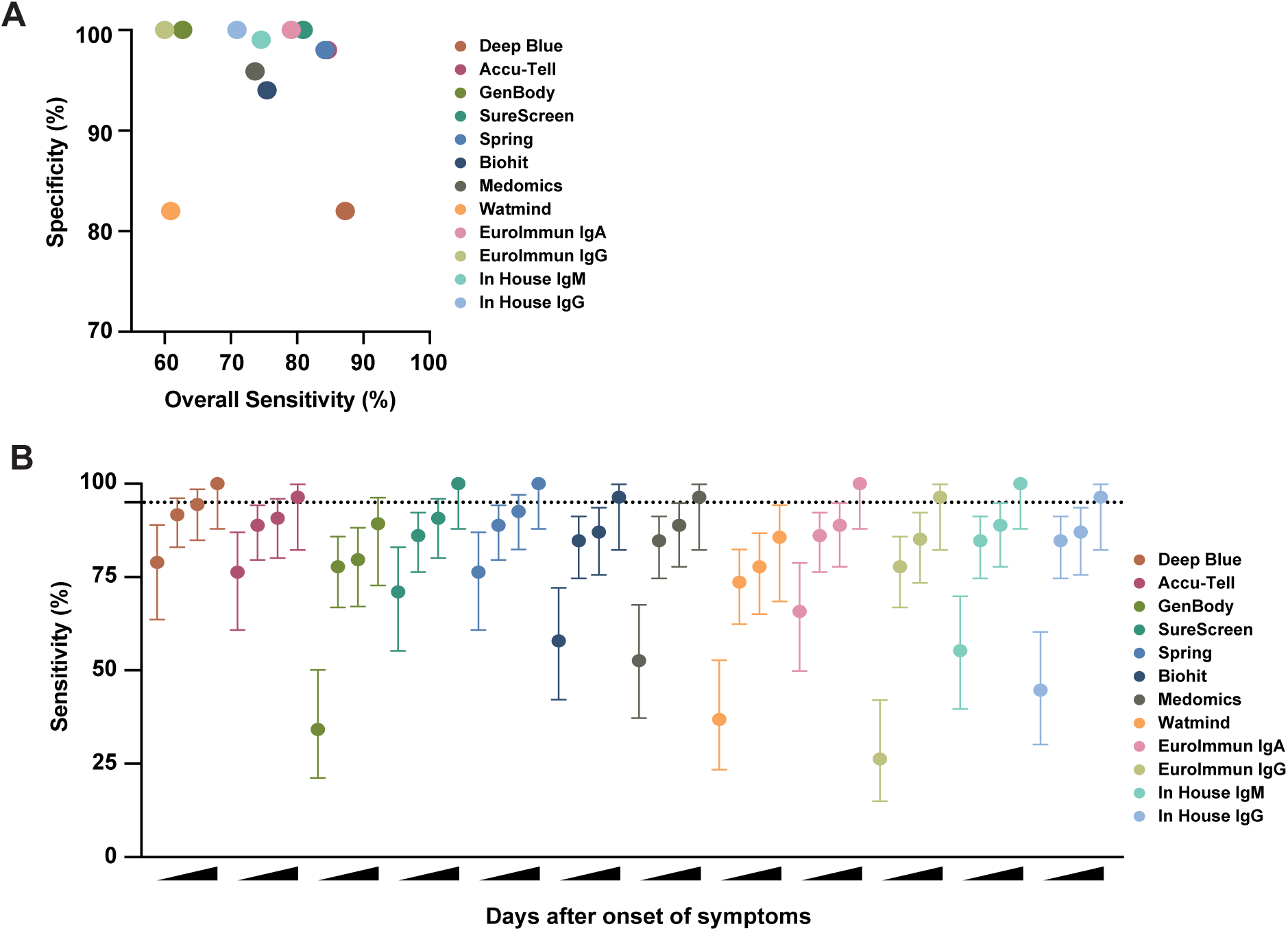
Sensitivity and specificity comparison of serological assays. (**A**) Specificity was determined for each serological assay using a panel of 50 pre-pandemic serum samples from March 2019. Overall sensitivity was determined for each serological assay, based on results for 110 serum samples from known SARS-CoV-2-positive individuals (shown in Figures 3 and 4). For the lateral flow assays, a positive result for IgM, IgG or both is considered positive. For the EUROIMMUN and in-house ELISAs, sensitivities for IgA, IgM and IgG are calculated separately. (**B**) Sensitivity and 95% confidence (dashed line) intervals were determined for each serological assay at increasing days POS. Results for each test were categorised according to whether the serum sample was from < 10, ≥10, ≥14, or ≥20 days POS.

### Association of antibody detection with days POS and severity of disease

Sample-by-sample analyses of multiple serological assays showed a trend for increasing detection of antibodies with increasing days POS (up to 30). We also observed that individuals with severe disease had a more readily detectable antibody response, particularly compared to those who had a brief hospital stay with minimal intervention. Samples were grouped according to days POS and clinical severity, and compared for anti-S IgM and IgG by in-house ELISA. Significant differences in antibody levels were observed with increases in both days and severity, although there was no association between days and severity themselves (**Figure 6A**). To assess the development of the antibody response in sequential samples from the same individual, and to evaluate the ability of LFIAs to detect nuances in antibody response, longitudinal samples (from individuals with disease severity scores of 4) were tested by one of the best performing LFIAs (SureScreen) and in-house ELISA (**Figure 6B** and **C**). A similar pattern of detection was seen for both types of assay, with IgM detectable earlier than IgG, and both tests showing consensus for the strength and timing of the response.

**Figure 6.**
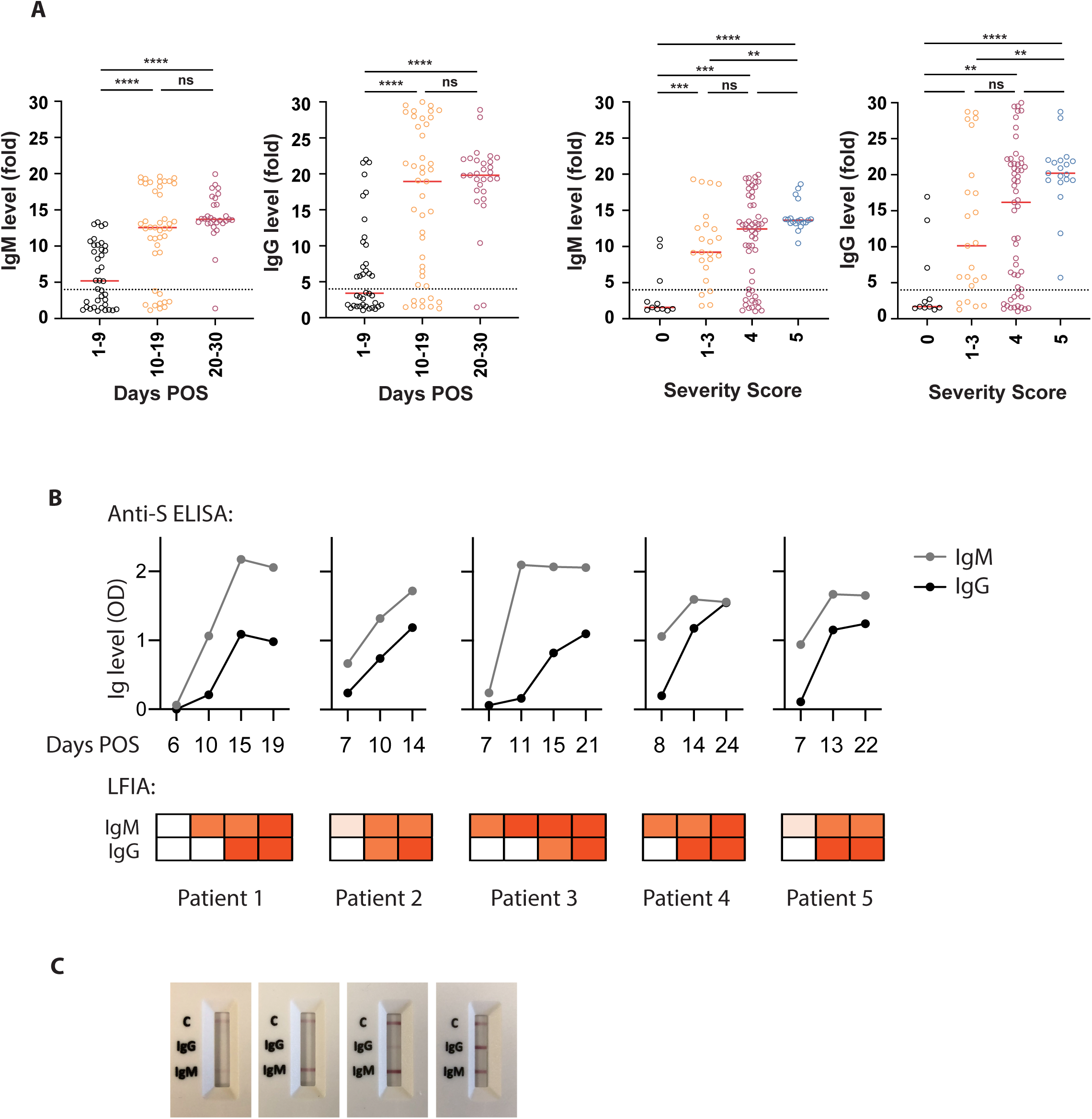
Antibody detection over increasing days POS and illness severity. (**A**) In-house ELISA results for anti-SARS-CoV-2 IgM and IgG were grouped by days POS and severity of illness, with 0 indicating mild illness (requiring no respiratory support) and 5 indicating critical (requiring ECMO) (see materials and methods for full classification). The 4-fold threshold for a positive result is shown as a dashed line on each graph. Median values are shown as red lines. (**B**) Sequential serum samples from five individuals were assayed for the development of anti-SARS-CoV-2 IgM and IgG. All five individuals were hospitalised with COVID-19 symptoms and confirmed positive for SARS-CoV-2 RNA by RT-PCR. A semi-quantitative in-house ELISA detecting anti-S antibodies (shown in graphs) and the SureScreen lateral flow assay (shown in heatmaps below the graphs) were compared for their detection of anti-SARS-CoV-2 in the same serum samples. Days POS are indicated for each sample. (C) Images of the lateral flow test results for patient 3 are shown.

## DISCUSSION

This study describes the development of six in-house ELISA configurations for the detection of IgM and IgG against SARS-CoV-2 S, RBD and N. Anti-S and –N IgG attained specificities of up to 99% – 100% when results for both targets are combined – and sensitivities of up to 96%. We used this semi-quantitative platform to cross-evaluate seven LFIAs, a chemiluminescent test and two commercial ELISAs. Importantly, the availability of sequential serum samples from patients admitted from the start of the outbreak under an existing ethics agreement for storage and analysis of surplus amounts of routinely collected clinical samples, enabled us to conduct this detailed study including examining assay sensitivity with respect to time POS.

Our analysis demonstrates a broad range of performance across the different platforms, with several commercial tests performing above 98% specificity. We found that all platforms showed highest sensitivity, with narrowest confidence limits, in samples taken 20 days POS, with most tests reaching a value of over 95% (**Figure 5B**). When all commercial tests were compared, Accu-Tell, SureScreen and Spring demonstrated highest sensitivity at earlier time points, while maintaining specificities of 98% or above. These tests also gave the best cross-assay agreements with each other and with the in-house ELISA. In the best-performing tests, we also observed that signal strength aligned with that seen by ELISA; this was further supported by the sequential signal increase seen in longitudinal samples from five individuals.

We approached this study with the intention that an unbiased and transparent comparison will be of broad value to the scientific and infection diagnostics communities, both in terms of naming and comparing the kits, and in the nature of the samples likely to be encountered in hospitals during a SARS-CoV-2 outbreak. Few studies have been published to date in which multiple LFIAs and ELISAs have been evaluated side-by-side with named kits [8–9]; and in those that have, only two tests overlap with our study, Deep Blue and Medomics [8]. Early reports stating that LFIAs have insufficient sensitivity may have been due to testing samples from mixed time points and disease severities, or tests that differ from those evaluated here [10].

The strength of our study is the head-to-head evaluation of multiple tests on identical serum samples. The sample set was not compiled retrospectively for the purpose of evaluation, but was part of an ongoing process to deploy a serological assay to broaden diagnostic capability at Guy’s and St Thomas’ Hospitals during the peak of the SARS-CoV-2 epidemic in London. These samples are therefore entirely representative of the type that will be encountered by hospital laboratories, and the challenges associated with variable time to seroconversion and errors in self-reporting onset of symptoms are real. The cross-evaluation of multiple tests enabled the identification of samples that were negative in every test performed, despite the fact that samples were taken at 10 days POS or later. While this could highlight a characteristic of COVID-19 where a subset of patients do not produce a detectable antibody response, we have evidence that in three cases (the two day 10 samples and sixth day 14) where later samples were available (47, 77 and 51 days POS, respectively; the only next available samples) both IgM and IgG antibodies were detected in several LFIAs. In another case (sample at day 23 POS), we uncovered a likely error in self-reported symptom onset and the sample could actually range anywhere between 2 and 11 days POS. It is important to note that all of the six unexpectedly negative samples were from COVID-19 cases classified as severity level 0.

While there is no current agreement on the relationship between disease severity and antibody titres (some show an association, some do not) [3, 8, 10–16], we observed a significant increase in detection of antibodies with increased severity of symptoms (**Figure 6A**). Importantly, this correlation is not explained by a concomitant increase in the days POS of the sample. Therefore, before deployment in situations where the pre-test prevalence is likely to be low, such as seroprevalence studies, out-patient assessment or pre-admission screening for operations, these assays will require further evaluation with known SARS-CoV-2 asymptomatic and ambulatory cases, alongside an extended set of pre-pandemic samples. This is a priority as countries navigate their way out of lockdown and move towards living with the ongoing threat of SARS-CoV-2, and seroprevalence studies will be important in the implementation and management of safe public health policies. To maintain the high specificity of our in-house ELISA in community cohorts where PCR status is unknown, we would recommend determining seropositivity based on IgG to both N and S. Sequential or alternate detection of IgM, IgA and IgG may also provide information on the history of infection. In the only IgA test that we evaluated (EUROIMMUN), specificity was high while also showing a strong signal in early samples and a good overall sensitivity. Detection of IgA in serum, and potentially even earlier by mucosal sampling, may be a useful diagnostic tool.

Next generation antibody tests may improve on those currently being trialled, but our results demonstrate that LFIAs may have utility in a hospital setting as of now, particularly if deployed where a rapid result could aide a clinical pathway or decision in real time, such as ward location or prioritisation of further diagnostics and follow up. The ease of use and affordability of the LFIAs weighs heavily in their favour, especially for potential in resource-poor settings or as point-of-care solutions in hospitals. Choosing the tests with the highest specificity will translate to confidence in a positive test result in the clinic; negative or borderline cases may be tested serially or used together with ELISA testing [17]. Combination testing, in parallel with RT-PCR, and serial or sequential testing, would provide diagnostic solutions to the delayed-onset syndromes such as PIMS that are increasingly being reported post-peak pandemic [5, 6]. A further consideration that should be given for healthcare service deployment in the hospital or community setting is consistency of use. Although the LFIAs are marketed as home testing kits, in our experience user assiduity is essential to their optimal performance, particularly when scoring borderline cases and considering need for two independent readers. There will also likely be need to evaluate alternative sources of blood collection particularly pin-prick collection, and even different samples such as saliva, all of which should be fully evaluated before deployment.

In summary, our study compares the performance of 10 commercially available platforms and several combinations of in-house methods for the detection of anti-SARS-CoV-2 antibodies in serum samples. Although LFIAs lack the semi-quantitative information provided by ELISA tests, they have a clear utility advantage over ELISA or chemiluminescence-based technologies. Shortlisted tests, combined with confirmatory reflex testing using our in-house ELISA, are now being taken forward into extended validations as part of a pilot clinical service at Guy’s and St Thomas’ Hospitals. These incremental steps keeping different technologies in scope, whilst multiple different use-cases are still being defined, will help determine the clinical utility and cost-effectiveness of COVID-19 serological testing in healthcare settings both in the hospital and the community.

## MATERIALS & METHODS

### Patient overview and sample origin

110 individual venous serum samples collected at St Thomas’ Hospital, London from 87 patients diagnosed as SARS-CoV-2 positive via real-time RT-PCR, were obtained for serological analysis. Samples ranged from 1 to 30 days after onset of self-reported symptoms. For the longitudinal study 17 serum samples (6–24 days after symptoms onset) were obtained from 5 patients (3–4 samples each) with confirmed COVID-19 diagnosis. Two patients overlapped between the longitudinal study and validation study meaning in total there are 90 unique patients between both studies. Patient information is given in **Table 1**. In addition, prepandemic serum samples from various collections were used as negative controls: 50 serum samples from 50 patients were obtained at St Thomas’ Hospital dating from March 2019; plasma samples collected from individuals 7 days following immunisation with an H1N1 vaccine (as part of the HIRD trial [18]) were obtained from the KCL Infectious Diseases Biobank.

### COVID-19 severity classification

Patients diagnosed with COVID-19 were classified as follows:

0 - asymptomatic OR no requirement for supplemental oxygen.

1 - requirement for supplemental oxygen (FiO_2_ < 0.4) for at least 12 hrs.

2 - requirement for supplemental oxygen (FiO_2_ ≥0.4) for at least 12 hrs.

3 - requirement for non-invasive ventilation (NIV)/ continuous positive airways pressure (CPAP) OR proning OR supplemental oxygen (FiO_2_ >0.6) for at least 12 hrs AND not a candidate for escalation above level 1 care.

4 - requirement for intubation and mechanical ventilation OR supplemental oxygen (FiO_2_ >0.8) AND peripheral oxygen saturations < 90% (with no history of type 2 respiratory failure (T2RF)) OR < 85% (with known T2RF) for at least 12 hrs.

5 - requirement for ECMO.

### ELISA protocol

All sera/plasma was heat-inactivated at 56°C for 30 mins before use in the in-house ELISA. High-binding ELISA plates (Corning, 3690) were coated with antigen (N, S or RBD) at 3 µg/mL (25 µL per well) in PBS, either overnight at 4°C or 2 hr at 37°C. Wells were washed with PBS-T (PBS with 0.05% Tween-20) and then blocked with 100 µL 5% milk in PBS-T for 1 hr at room temperature. Wells were emptied and sera and plasma diluted at 1:50 and 1:25 respectively in milk were added and incubated for 2 hr at room temperature. Control reagents included CR3009 (2 µg/mL), CR3022 (0.2 µg/mL), negative control plasma (1:25 dilution), positive control plasma (1:50) and blank wells. Wells were washed with PBS-T. Secondary antibody was added and incubated for 1 hr at room temperature. IgM was detected using Goat-antihuman-IgM-HRP (1:1,000) (Sigma: A6907) and IgG was detected using Goat-anti-human-Fc-AP (1:1,000) (Jackson: 109–055–043-JIR). Wells were washed with PBS-T and either AP substrate (Sigma) was added and read at 405 nm (AP) or 1-step TMB substrate (Thermo Scientific) was added and quenched with 0.5 M H_2_S0_4_ before reading at 450 nm (HRP).

### Protein expression

N protein was obtained from the James lab at LMB, Cambridge. The N protein used is a truncated construct of the SARS-CoV-2 N protein comprising residues 48–365 (both ordered domains with the native linker) with an N terminal uncleavable hexahistidine tag. N was expressed in *E. Coli* using autoinducing media for 7h at 37°C and purified using immobilised metal affinity chromatography (IMAC), size exclusion and heparin chromatography.

S protein consists of a pre-fusion S ectodomain residues 1–1138 with proline substitutions at amino acid positions 986 and 987, a GGGG substitution at the furin cleavage site (amino acids 682–685) and an N terminal T4 trimerisation domain followed by a Strep-tag II [19]. The protein was expressed in 1 L HEK-293F cells (Invitrogen) grown in suspension at a density of 1.5 million cells/mL. The culture was transfected with 325 µg of DNA using PEI-Max (1 mg/mL, Polysciences) at a 1:3 ratio. Supernatant was harvested after 7 days and purified using StrepTactinXT Superflow high capacity 50% suspension according to the manufacturer’s protocol by gravity flow (IBA Life Sciences).

The RBD plasmid was obtained from Florian Krammer at Mount Sinai University [20]. Here the natural N-terminal signal peptide of S is fused to the RBD sequence (319 to 541) and joined to a C-terminal hexahistidine tag. This protein was expressed in 500 mL HEK-293F cells (Invitrogen) at a density of 1.5 million cells/mL. The culture was transfected with 1000 µg of DNA using PEI-Max (1 mg/mL, Polysciences) at a 1:3 ratio. Supernatant was harvested after 7 days and purified using Ni-NTA agarose beads.

### Lateral Flow Immunoassays (LFIA)

We tested seven point-of-care colloidal-gold-based LFIAs detecting IgG and IgM antibodies against SARS-CoV-2. With the exception of Medomics, all LFIAs were CE IVD marked. Target antigens were undisclosed proprietary information, but for several of them were known to be S. LFIAs were run according to manufacturer’s instructions. Typically, 10–20 µl of serum was added to the LFIA membrane start point, followed by 1–3 drops of supplied buffer. Kits were run at room temperature for 10 minutes and then immediately scored using a 4-point scale (negative, borderline, positive, strong positive) for both IgM and IgG. Scoring was performed independently by two individuals.

| **LFIA** | **Distributor/Manufacturer** | **Reference** | **Lot No.** |
| --- | --- | --- | --- |
| Accu-Tell COVID-19 IgG/IgM Cassette | AccuBiotech Co., Ltd. | ABT-IDT-B352 | #2020031701 |
| COVID-19 (SARS-CoV-2) Antibody Test Kit | Anhui DeepBlue Medical Technology Co., Ltd. | N/A | #20200305 |
| SARS-CoV-2 IgM/IgG ANTIBODY TEST KIT | Biohit Healthcare Co., Ltd. | N/A | SA200401 |
| GenBody COVID-19 IgM/IgG | GenBody Inc. | COVI025 | FJFN25201 |
| COVID-19 Spring IgM/IgG Rapid Test Cassette | Spring Healthcare Services AG | ERCSSO5310 | N01G26T |
| COVID-19 IgG/IgM Rapid Test Cassette | SureScreen Diagnostics Co., Ltd. | COVID19C | COV20030045 |
| Rapid IgM-IgG Combined Antibody Test Kit for SARS-CoV-2 | Jiangsu Medomics Medical Technology Co. Ltd. | N/A | #2020050305 |

### Chemiluminescence-based Immunoassay

The SARS-CoV-2 Ab Diagnostic Test Kit (Shenzen Watmind Medical Co., Ltd.) detecting total antibody against SARS-CoV-2 was run on the Chemical Luminescence Immunity Analyzer MF02 (Shenzen Watmind Medical Co., Ltd). The platform was calibrated with a supplied control cartridge daily prior to testing. Panel samples were analysed according to manufacturer’s instructions. Results equal to and below 1.0 AU (arbitrary units) /ml were negative, scores above 1.0 AU/ml were deemed positive. For comparison to other immunoassays, scores between 1< 10 AU/ml were deemed positive, scores > 10 AU/ml were deemed a strong positive.

### Enzyme-linked Immunosorbent Assay (ELISA)

Two commercial ELISAs detecting IgA (EI 2606–9601 A) or IgG (EI 2606–9601 G) antibodies to the SARS-CoV-2 S1 protein were obtained from EUROIMMUN Medizinische Labordiagnostika AG. Serum samples were tested according to manufacturer’s instructions. Absorbance was measured on a Multiskan FC (SkanIt Software 5.0 for Microplate Readers RE. ver. 5.0.0.42) at a wavelength of 450 nm with a reference wavelength of 630 nm.

### Statistical Analysis

Expected binomial exact 95% confidence intervals were calculated on Prism 8.0 using Wilson/Brown statistical analysis.

**Supplementary Figure A.**
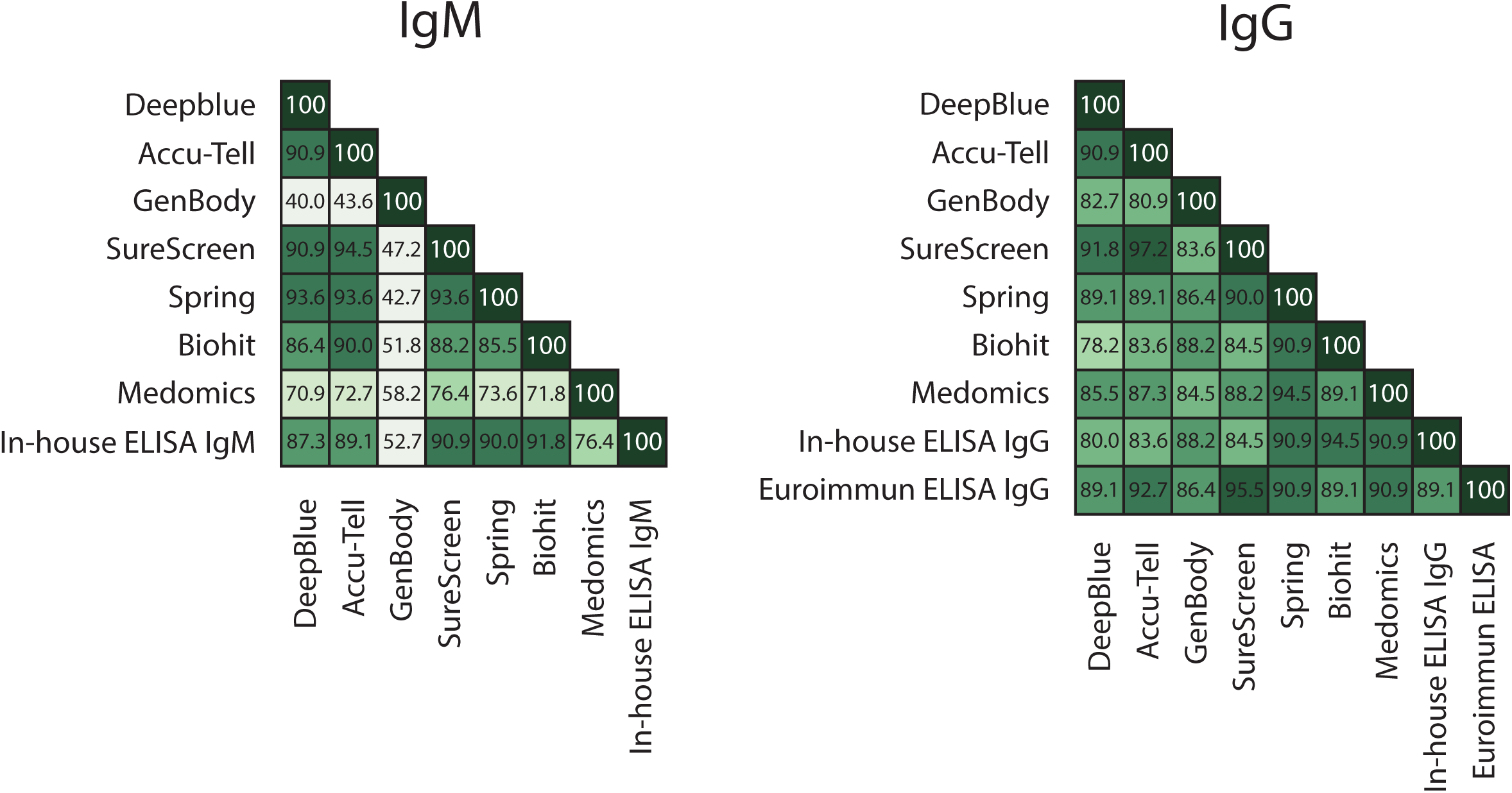
Serological assays were compared for IgM or IgG (left and right panels, respectively), and the percentage agreement between each of the samples in the assays is represented within each box.

**Supplementary Figure B.**
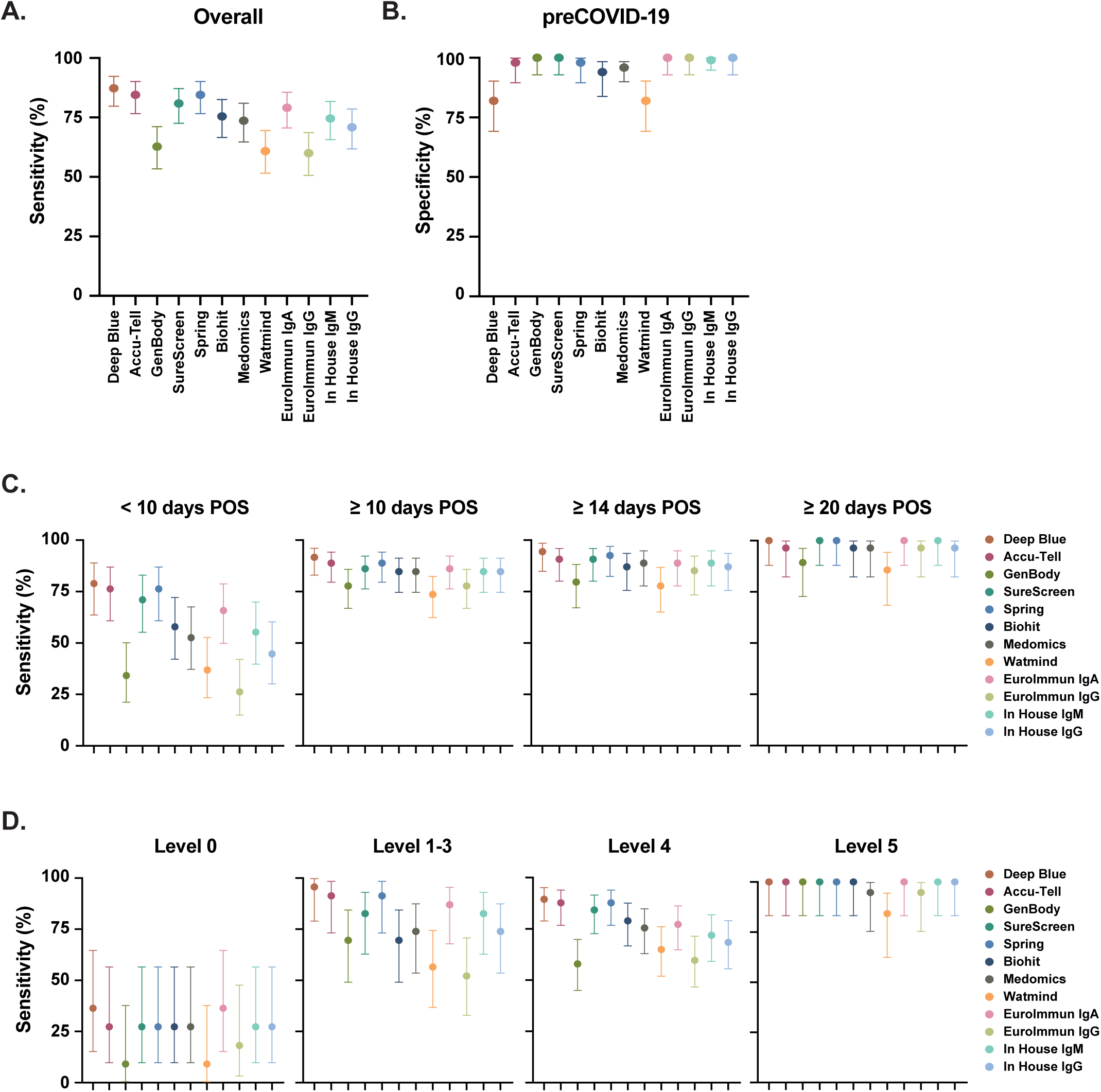
Overall sensitivity (**A**) and specificity (**B**) were determined for each serological assay as in Figure 5. Sensitivity was determined for each serological assay at increasing days POS (**C**), or severity of illness (**D**). Results for each test were either categorised according to whether the serum sample was from < 10, ≥10, ≥14, or ≥20 days POS, or severity of illness, with 0 indicating mild illness (requiring no respiratory support) and 5 indicating critical (requiring ECMO) (see Materials and Methods for full classification). 95% confidence intervals are shown for each assay in all panels (Wilson/Brown expected binomial).

### Funding

The research and the King’s College London Infectious Diseases Biobank were supported by the Department of Health via a National Institute for Health Research comprehensive Biomedical Research Centre award to Guy’s and St. Thomas’ NHS Foundation Trust in partnership with King’s College London and King’s College Hospital NHS Foundation Trust.

AWS and CG were supported by the MRC-KCL Doctoral Training Partnership in Biomedical Sciences (MR/N013700/1).

GB was supported by the Wellcome Trust (106223/Z/14/Z to MHM).

SA was supported by an MRC-KCL Doctoral Training Partnership in Biomedical Sciences industrial Collaborative Award in Science & Engineering (iCASE) in partnership with Orchard Therapeutics (MR/R015643/1).

NK was supported by the Medical Research Council (MR/S023747/1 to MHM).

SP, HDW and SJDN were supported by a Wellcome Trust Senior Fellowship (WT098049AIA).

King’s Together Rapid COVID-19 Call awards to MHM, KJD, SJDN and RMN.

Fondation Dormeur, Vaduz for funding equipment (KJD).

MRC Discovery Award MC/PC/15068 to SJDN, KJD and MHM.

Development of SARS-CoV-2 reagents (RBD) was partially supported by the NIAID Centers of Excellence for Influenza Research and Surveillance (CEIRS) contract HHSN272201400008C.

### Ethics

For the St Thomas’ Hospital samples, surplus serum was retrieved from the routine biochemistry laboratory at point of discard, and then aliquoted, stored and linked with a limited clinical dataset by the direct care team, before anonymisation under an existing ethics framework (REC reference 18/NW/0584) and with expedited R&D approval. Serum/plasma samples used as negative controls in the in-house ELISA development were obtained from the KCL Infectious Disease Biobank (KD1-220518), UCLH (11/LO/0421 IRAS:43993) and The Royal Free Hospital (11/WA/0077, ref. NC2016.002).

## Data Availability

Aggregate anonymised data was used in this study and is not available for distribution outside the host organisations.

## Acknowledgements

The work was supported by gifts from Peking University donors and Anhui Deep Blue company. Thank you to Dr Terry Wong for his support in acquiring test kits. We also thank the following sources for donation of test kits: the manufacturers of Spring, Biohit, Genbody, Medomics and Watmind. Thank you to SureScreen Diagnostics for their engagement and technical assistance.

Thank you to Bindi Patel, Nicola Varatharajah, Abayomi Fatola and Amelia Moore for laboratory assistance.

Thank you to Florian Krammer for provision of the RBD expression plasmid, and Leo James and Leo Kiss for the provision of purified N protein.

We thank King’s College London Infectious Diseases Biobank for provision of pre-COVID-19 vaccine samples, all patients and control volunteers who participated in this study and to all clinical staff who helped with recruitment, including those working with the TAPb project at The Royal Free Hospital. We thank the Maini Lab at the Division of Infection and Immunity for providing pre-Covid19 pandemic healthy control samples.

We are extremely grateful to all patients and staff at St Thomas’ Hospital who participated in this study.

---

**Supplemental Table 1.** Sensitivity and Specificity of in-house ELISAs during development phase, with 95% CI.

|  | Specificity |  |  |  | Sensitivity |  |  |  |
| --- | --- | --- | --- | --- | --- | --- | --- | --- |
|  | Total N | Negatives | % | 95% CI | Total N | Positives | % | 95% CI |
| **In-house IgM - N** | 320 | 200 | 62.50 | 57.08 to 67.63 | 24 | 22 | 91.67 | 74.15 - 98.52 |
| **In-house IgG - N** | 320 | 271 | 84.69 | 80.33 to 88.22 | 24 | 24 | 100.00 | 86.20 - 100.0 |
| **In-house IgM - S** | 320 | 291 | 90.94 | 87.29 to 93.62 | 24 | 24 | 100.00 | 86.20 - 100.0 |
| **In-house IgG - S** | 320 | 316 | 98.75 | 96.83 to 99.51 | 24 | 24 | 100.00 | 86.20 - 100.0 |
| **In-house IgM - RBD** | 320 | 305 | 95.31 | 92.41 to 97.14 | 24 | 24 | 100.00 | 86.20 - 100.0 |
| **In-house IgG - RBD** | 320 | 319 | 99.69 | 98.25 to 99.98 | 24 | 24 | 100.00 | 86.20 - 100.0 |
| **In-house IgM N+S** | 320 | 301 | 94.06 | 90.91 to 96.17 | 24 | 22 | 91.67 | 74.15 - 98.52 |
| **In-house IgG N+S** | 320 | 320 | 100.00 | 98.81 to 100.0 | 24 | 24 | 100.00 | 86.20 - 100.0 |
| **In-house IgM N+RBD** | 320 | 309 | 96.56 | 93.95 to 98.07 | 24 | 22 | 91.67 | 74.15 - 98.52 |
| **In-house IgG N+RBD** | 320 | 320 | 100.00 | 98.81 to 100.0 | 24 | 24 | 100.00 | 86.20 - 100.0 |

**Supplemental Table 2.**
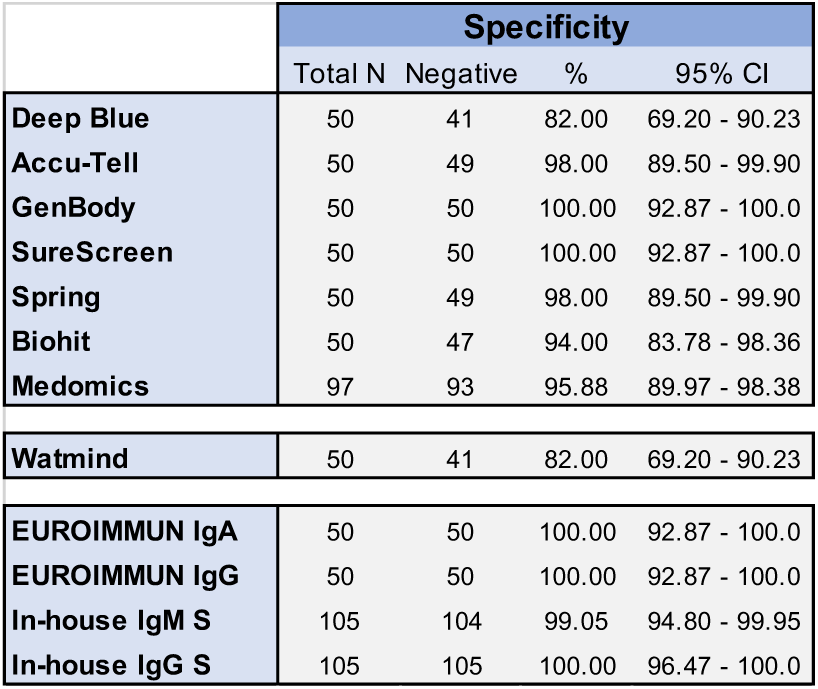
Specificity of immunoassays platforms determined in blood donor plasma specimens collected on March 2019.

|  | Specificity |  |  |  |
| --- | --- | --- | --- | --- |
|  | Total N | Negative | % | 95% CI |
| Deep Blue | 50 | 41 | 82.00 | 69.20 - 90.23 |
| Accu-Tell | 50 | 49 | 98.00 | 89.50 - 99.90 |
| GenBody | 50 | 50 | 100.00 | 92.87 - 100.0 |
| SureScreen | 50 | 50 | 100.00 | 92.87 - 100.0 |
| Spring | 50 | 49 | 98.00 | 89.50 - 99.90 |
| Biohit | 50 | 47 | 94.00 | 83.78 - 98.36 |
| Medomics | 97 | 93 | 95.88 | 89.97 - 98.38 |
| Watmind | 50 | 41 | 82.00 | 69.20 - 90.23 |
| EUROIMMUN IgA | 50 | 50 | 100.00 | 92.87 - 100.0 |
| EUROIMMUN IgG | 50 | 50 | 100.00 | 92.87 - 100.0 |
| In-house IgM S | 105 | 104 | 99.05 | 94.80 - 99.95 |
| In-house IgG S | 105 | 105 | 100.00 | 96.47 - 100.0 |

**Supplemental Table 3.** Sensitivity of different platforms determined on specimens from patients with positive SARS-CoV-2 RT-PCR by days since symptom onset.

|  | OVERALL |  |  |  | < 10 days |  |  |  | ≥ 10 days |  |  |  | ≥ 14 days |  |  |  | ≥ 20 days |  |  |  |
| --- | --- | --- | --- | --- | --- | --- | --- | --- | --- | --- | --- | --- | --- | --- | --- | --- | --- | --- | --- | --- |
|  | Total N | Positive | % | 95% CI | Total N | Positive | % | 95% CI | Total N | Positive | % | 95% CI | Total N | Positive | % | 95% CI | Total N | Positive | % | 95% CI |
| Deep Blue | 110 | 96 | 87.27 | 79.76 - 92.27 | 38 | 30 | 78.95 | 63.65 - 88.93 | 72 | 66 | 91.67 | 82.99 - 96.12 | 54 | 51 | 94.44 | 84.89 - 98.49 | 28 | 28 | 100.00 | 87.94 - 100.0 |
| Accu-Tell | 110 | 93 | 84.55 | 76.64 - 90.12 | 38 | 29 | 76.32 | 60.79 - 87.01 | 72 | 64 | 88.89 | 79.58 - 94.26 | 54 | 49 | 90.74 | 80.09 - 95.98 | 28 | 27 | 96.43 | 82.29 - 99.82 |
| GenBody | 110 | 69 | 62.73 | 53.41 - 71.19 | 38 | 13 | 34.21 | 21.21 - 50.11 | 72 | 56 | 77.78 | 66.91 - 85.83 | 54 | 43 | 79.63 | 67.10 - 88.23 | 28 | 25 | 89.29 | 72.80 - 96.29 |
| SureScreen | 110 | 89 | 80.91 | 72.57 - 87.16 | 38 | 27 | 71.05 | 55.24 - 83.00 | 72 | 62 | 86.11 | 76.29 - 92.28 | 54 | 49 | 90.74 | 80.09 - 95.98 | 28 | 28 | 100.00 | 87.94 - 100.0 |
| Spring | 110 | 93 | 84.55 | 76.64 - 90.12 | 38 | 29 | 76.32 | 60.79 - 87.01 | 72 | 64 | 88.89 | 79.58 - 94.26 | 54 | 50 | 92.59 | 82.45 - 97.08 | 28 | 28 | 100.00 | 87.94 - 100.0 |
| Biohit | 110 | 83 | 75.45 | 66.64 - 82.55 | 38 | 22 | 57.89 | 42.19 - 72.15 | 72 | 61 | 84.72 | 74.68 - 91.25 | 54 | 47 | 87.04 | 75.58 - 93.58 | 28 | 27 | 96.43 | 82.29 - 99.82 |
| Medomics | 110 | 81 | 73.64 | 64.71 - 80.97 | 38 | 20 | 52.63 | 37.26 - 67.52 | 72 | 61 | 84.72 | 74.68 - 91.25 | 54 | 48 | 88.89 | 77.81 - 94.81 | 28 | 27 | 96.43 | 82.29 - 99.82 |
| Watmind | 110 | 67 | 60.91 | 51.57 - 69.51 | 38 | 14 | 36.84 | 23.38 - 52.72 | 72 | 53 | 73.61 | 62.42 - 82.41 | 54 | 42 | 77.78 | 65.06 - 86.80 | 28 | 24 | 85.71 | 68.51 - 94.30 |
| EUROIMMUN IgA | 110 | 87 | 79.09 | 70.57 - 85.64 | 38 | 25 | 65.79 | 49.89 - 78.79 | 72 | 62 | 86.11 | 76.29 - 92.28 | 54 | 48 | 88.89 | 77.81 - 94.81 | 28 | 28 | 100.00 | 87.94 - 100.0 |
| EUROIMMUN IgG | 110 | 66 | 60.00 | 50.66 - 68.67 | 38 | 10 | 26.32 | 14.97 - 42.01 | 72 | 56 | 77.78 | 66.91 - 85.83 | 54 | 46 | 85.19 | 73.40 - 92.30 | 28 | 27 | 96.43 | 82.29 - 99.82 |
| In-house IgM S | 110 | 82 | 74.55 | 65.67 - 81.76 | 38 | 21 | 55.26 | 39.71 - 69.85 | 72 | 61 | 84.72 | 74.68 - 91.25 | 54 | 48 | 88.89 | 77.81 - 94.81 | 28 | 28 | 100.00 | 87.94 - 100.0 |
| In-house IgG S | 110 | 81 | 70.91 | 61.83 - 78.58 | 38 | 20 | 52.63 | 37.26 - 67.52 | 72 | 61 | 84.72 | 74.68 - 91.25 | 54 | 47 | 87.04 | 75.58 - 93.58 | 28 | 27 | 96.43 | 82.29 - 99.82 |

**Supplemental Table 4.** Sensitivity of different platforms determined on specimens from patients with positive SARS-CoV-2 RT-PCR by disease severity.

|  | Level 0 |  |  |  | Levels 1-3 |  |  |  | Level 4 |  |  |  | Level 5 |  |  |  |
| --- | --- | --- | --- | --- | --- | --- | --- | --- | --- | --- | --- | --- | --- | --- | --- | --- |
|  | Total N | Positive | % | 95% CI | Total N | Positive | % | 95% CI | Total N | Positive | % | 95% CI | Total N | Positive | % | 95% CI |
| **Deep Blue** | 11 | 4 | 36.36 | 15.17 - 64.62 | 23 | 22 | 95.65 | 79.01 - 99.78 | 57 | 51 | 89.47 | 78.88 - 95.09 | 19 | 19 | 100.00 | 83.18 - 100.0 |
| **Accu-Tell** | 11 | 3 | 27.27 | 9.75 - 56.56 | 23 | 21 | 91.30 | 73.20 - 98.45 | 57 | 50 | 87.72 | 76.75 - 93.92 | 19 | 19 | 100.00 | 83.18 - 100.0 |
| **GenBody** | 11 | 1 | 9.09 | 0.46 - 37.74 | 23 | 16 | 69.57 | 49.13 - 84.40 | 57 | 33 | 57.89 | 44.98 - 69.81 | 19 | 19 | 100.00 | 83.18 - 100.0 |
| **SureScreen** | 11 | 3 | 27.27 | 9.75 - 56.56 | 23 | 19 | 82.61 | 62.86 - 93.02 | 57 | 48 | 84.21 | 72.64 - 91.46 | 19 | 19 | 100.00 | 83.18 - 100.0 |
| **Spring** | 11 | 3 | 27.27 | 9.75 - 56.56 | 23 | 21 | 91.30 | 73.20 - 98.45 | 57 | 50 | 87.72 | 76.75 - 93.92 | 19 | 19 | 100.00 | 83.18 - 100.0 |
| **Biohit** | 11 | 3 | 27.27 | 9.75 - 56.56 | 23 | 16 | 69.57 | 49.13 - 84.40 | 57 | 45 | 78.95 | 66.71 - 87.53 | 19 | 19 | 100.00 | 83.18 - 100.0 |
| **Medomics** | 11 | 3 | 27.27 | 9.75 - 56.56 | 23 | 17 | 73.91 | 53.53 - 87.45 | 57 | 43 | 75.44 | 62.90 - 84.77 | 19 | 18 | 94.74 | 75.36 - 99.73 |
| **Watmind** | 11 | 1 | 9.09 | 0.46 - 37.74 | 23 | 13 | 56.52 | 36.81 - 74.37 | 57 | 37 | 64.91 | 51.94 - 76.00 | 19 | 16 | 84.21 | 62.43 - 94.48 |
| **EUROIMMUN IgA** | 11 | 4 | 36.36 | 15.17 - 64.62 | 23 | 20 | 86.96 | 67.87 - 95.46 | 57 | 44 | 77.19 | 64.79 - 86.16 | 19 | 19 | 100.00 | 83.18 - 100.0 |
| **EUROIMMUN IgG** | 11 | 2 | 18.18 | 3.23 - 47.70 | 23 | 12 | 52.17 | 32.96 - 70.76 | 57 | 34 | 59.65 | 46.70 - 71.38 | 19 | 18 | 94.74 | 75.36 - 99.73 |
| **In-house IgM S** | 11 | 3 | 27.27 | 9.75 - 56.56 | 23 | 19 | 82.61 | 62.86 - 93.02 | 57 | 41 | 71.93 | 59.17 - 81.92 | 19 | 19 | 100.00 | 83.18 - 100.0 |
| **In-house IgG S** | 11 | 3 | 27.27 | 9.75 - 56.56 | 23 | 18 | 78.26 | 58.10 - 90.34 | 57 | 41 | 71.93 | 59.17 - 81.92 | 19 | 19 | 100.00 | 83.18 - 100.0 |

## References

1. NHS. Guidance and Standard Operating Procedure COVID-19 Virus Testing in NHS Laboratories (https://www.rcpath.org/uploads/assets/90111431-8aca-4614-b06633d07e2a3dd9/Guidance-and-SOP-COVID-19-Testing-NHS-Laboratories.pdf)

2. Centers for Disease Control and Prevention. Interim Guidelines for Collecting, Handling, and Testing Clinical Specimens for COVID-19 (https://www.cdc.gov/coronavirus/2019-nCoV/lab/guidelines-clinicalspecimens.html)

3. To KK, Tsang OT, Leung WS, Tam AR, Wu TC, Lung DC, et al. Temporal profiles of viral load in posterior oropharyngeal saliva samples and serum antibody responses during infection by SARS-CoV-2: an observational cohort study. Lancet Infect Dis. 2020;20(5):565–74.

4. Zou L, Ruan F, Huang M, Liang L, Huang H, Hong Z, et al. SARS-CoV-2 Viral Load in Upper Respiratory Specimens of Infected Patients. N Engl J Med. 2020;382(12):1177–9.

5. Verdoni L, Mazza A, Gervasoni A, Martelli L, Ruggeri M, Ciuffreda M, et al. An outbreak of severe Kawasaki-like disease at the Italian epicentre of the SARS-CoV-2 epidemic: an observational cohort study. Lancet. 2020.

6. European Centre for Dieases Prevention and Control. Paediatric inflammatory multisystem syndrome and SARS-CoV-2 infection in children (https://www.ecdc.europa.eu/en/publications-data/paediatric-inflammatorymultisystem-syndrome-and-sars-cov-2-rapid-risk-assessment)

7. Randolph HE, Barreiro LB. Herd Immunity: Understanding COVID-19. Immunity. 2020;52(5):737–41.

8. Whitman JD, Hiatt J, Mowery CT, Shy BR, Yu R, Yamamoto TN, et al. Test performance evaluation of SARS-CoV-2 serological assays. medRxiv 2020.04.25.20074856 [Preprint]. 2020 [cited 2020 May 29]. Available from: https://doi.org/10.1101/2020.04.25.20074856.

9. Lassaunière R, Frische A, Harboe ZB, Nielsen ACY, Fomsgaard A, Krogfelt, Jørgensen CS. Evaluation of nine commercial SARS-CoV-2 immunoassays. medRxiv 2020.04.09.20056325 [Preprint]. 2020 [cited 2020 May 29]. Available from: https://doi.org/10.1101/2020.04.09.20056325.

10. Adams E, Ainsworth M, Anand R, Andersson MI, Auckland K, et al. Antibody testing for COVID-19: A report from the National COVID Scientific Advisory Panel. medRxiv 2020.04.15.20066407 [Preprint]. 2020 [cited 2020 May 29]. Available from: https://doi.org/10.1101/2020.04.15.20066407.

11. Zhao J, Yuan Q, Wang H, Liu W, Liao X, Su Y, et al. Antibody responses to SARS-CoV-2 in patients of novel coronavirus disease 2019. Clin Infect Dis. 2020.

12. Tan W, Lu Y, Wang J, Dan Y, Tan Z, He, X, et al. Viral Kinetics and Antibody Responses in Patients with COVID-19. medRxiv 2020.03.24.20042382 [Preprint]. 2020 [cited 2020 May 29]. Available from: https://doi.org/10.1101/2020.03.24.20042382.

13. Wu F, Wang A, Liu M, Wang Q, Chen J, Xia S, et al. Neutralizing antibody responses to SARS-CoV-2 in a COVID-19 recovered patient cohort and their implications. medRxiv 2020.03.30.20047365 [Preprint]. 2020 [cited 2020 May 29]. Available from: https://doi.org/10.1101/2020.03.30.20047365.

14. Long QX, Liu BZ, Deng HJ, Wu GC, Deng K, Chen YK, et al. Antibody responses to SARSCoV-2 in patients with COVID-19. Nat Med. 2020.

15. Sun B, Feng Y, Mo X, Zheng P, Wang Q, Li P, et al. Kinetics of SARS-CoV-2 specific IgM and IgG responses in COVID-19 patients. Emerg Microbes Infect. 2020;9(1):940–8.

16. Lon Q, Deng H, Chen J, Hu J, Liu B, Liao P, et al. Antibody responses to SARS-CoV-2 in COVID-19 patients: the perspective application of serological tests in clinical practice. medRxiv 2020.03.18.20038018 [Preprint]. 2020 [cited 2020 May 29]. Available from: https://doi.org/10.1101/2020.03.18.20038018.

17. Egners W. Recommendations for verification and validation methodology and sample sets for evaluation of assays for SARS-CoV-2 (COVID-19). https://www.rcpath.org/uploads/assets/541a4523-6058-4424-81c119dd2ab0febb/Verification-validation-of-sample-sets-assays-SARS-CoV-2.pdf

18. Sobolev O, Binda E, O’Farrell S, Lorenc A, Pradines J, Huang Y, Duffner J, Schulz R, Cason J, Zambon M, Malim MH, Peakman M, Cope A, Capila I, Kaundinya GV and Hayday AC. Adjuvented influenza-H1N1 vaccination reveals lymphoid signatures of age-dependent early responses and of clinical adverse events. Nat Immunol. 2016 Feb; 17(2): 204–213.

19. Brouwer PJM, Caniels TG, van der Straten K, Snitselaar JL, et al. Potent neutralizing antibodies from COVID-19 patients define multiple targets of vulnerability. BioRxiv 2020.05.12.088716. Available from: https://www.biorxiv.org/content/10.1101/2020.05.12.088716v1

20. Amanat F, Stadlbauer D et al. A serological assay to detect SARS-CoV-2 seroconversion in humans. Nat Med. 2020 s41591-020-0913-5.

